# A tissue multiplexing approach to maximise spatial transcriptomics output from precious pathology specimens

**DOI:** 10.1101/2025.02.06.25321364

**Authors:** Yung-Ching Kao, Katie J. Lee, Chenhao Zhou, Andrew Causer, Harald Oey, Kiarash Khosrotehrani, Blake O’Brien, Angus Collins, Quan Nguyen, H. Peter Soyer, Mitchell S. Stark

**Affiliations:** Frazer Institute, The University of Queensland, Dermatology Research Centre, Brisbane, QLD 4102, Australia; Institute for Molecular Bioscience, the University of Queensland, Saint Lucia, Queensland, Australia; Department of Dermatology, Princess Alexandra Hospital, Brisbane, Queensland, Australia; Sullivan Nicolaides Pathology, Brisbane, Australia; QIMR Berghofer Medical Research Institute, Herston, QLD, Australia

## Abstract

Since diagnostic specimens retrieved from pathology cannot be altered (except for sectioning) for ethicolegal reasons, we describe a method that combines multiple patient specimens, cut from individual blocks, allowing the measurement of several tissues in one capture array. This results in a cost-effective means to increase sample size to permit statistically robust data outputs and to fast-track clinical implementation. Importantly, our tissue multiplexing method is compatible with the standard 10X Genomics supported protocols. We have used skin biopsies in this instance as this is our primary area of research, but this method can be extended to any tissue of interest.

## Main text

Spatial transcriptomics (ST) is a cutting-edge technology that enables spatially resolved whole transcriptome detection across fresh frozen and formalin-fixed paraffin-embedded (FFPE) tissue sections (1, 2). Through ST, positional context of cell populations and cell-cell interactions can be revealed which is essential for improving our understanding of disease progression and treatment efficacy (3). Common limitations to the ST workflow include a relatively small capture area for tissue placement, long processing time of individual samples combined with the high financial costs for reagents and sequencing.

In most cases, if diagnostic tissue blocks can be retrieved from pathology for research purposes, these cannot be used to generate a tissue microarray or removed in its entirety from the original block due to medico-legal and ethical constraints. Moreover, each paraffin block often contains multiple tissues generated from the same biopsy, and upon haematoxylin and eosin (H&E) review of sections, not all tissues contain the area of interest. The power of the Visium CytAssist technology is the ability to work with pre-sectioned tissue slides, by covering the area of interest through a moveable gasket. This is particularly useful when only tissue sections are available from pathology, or if a smaller region of a large tissue biopsy is the focal point. Here, using skin biopsies as an example, we describe a modified Visium CytAssist (10xGenomics) tissue preparation workflow that maximises the capture area (6.5 × 6.5mm or 11 × 11mm window) covered by tissue sections, as a cost-effective strategy to increase sample size.

Skin biopsies (including lesions suspicious for melanoma or keratinocyte cancer) excised for diagnostic purposes vary in size (~2-10mm^3^ to multiple cm^3^), and previous ST has highlighted that most genes are detected in the epidermis and dermo-epidermal junction, and are usually sparse in the dermis or subcutaneous tissue (4). As such, if only one tissue section per slide is used, this may only cover ~10% of the capture area which is an inefficient use of these powerful tools. This can be addressed by combining many tissue specimens/regions of interest, cut from multiple tissue sections, into a single tissue slide (**Figure 1a**). The tissues are placed within the movable gasket window region to maximise transfer of multiple tissues (in this instance n=17) to one Visium capture window.

**Figure 1.**
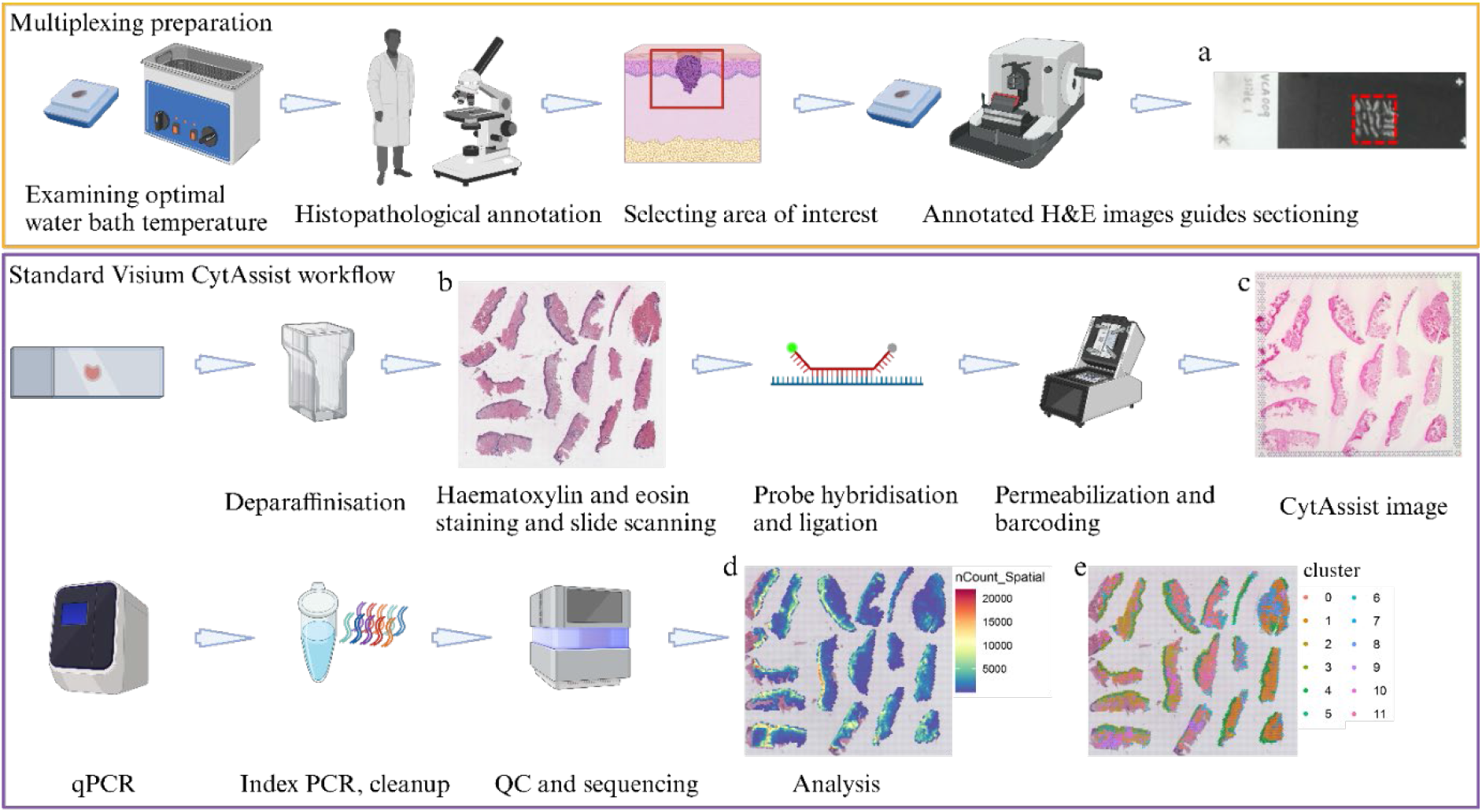
Illustration of multiplexing 17 skin tissues in one 11mm x 11mm Visium capture area. Multiplexing tissues for Visium CytAssist workflow requires optimising the water bath temperature for each sample, selecting the area of interest and isolating the area of interest by performing dissection on a 4µm section with a scalpel blade (on a microtome stage) or forceps (in a water bath). After incubation in a water bath, the isolated area of interest is placed as closely as possible within a pre-drawn 11 × 11 mm designated square (outlined in red) on a positively charged tissue slide (**a)**. This step is repeated for each sample until the pre-drawn 11 × 11 mm area is covered entirely. Compatible tissue slides and allowable area for CytAssist assay are detailed in the sample preparation protocol CG000520 (10x Genomics). Once the tissue slides containing the FFPE tissue sections are dried overnight in a desiccator, the tissues are stained with haematoxylin and eosin (H&E) and scanned (**b)** following protocol CG000520 (10X Genomics). The H&E-stained slide is then subjected to destaining, probe hybridisation, probe ligation as per protocol CG000495 (10X Genomics). Once the processed tissue slide is ready for CytAssist-enabled transfer, the slide is stained again with eosin to mark the tissue regions to match the Visium CytAssist capture area. As part of the CytAssist-enabled transfer of the gene expression probes released from the tissues and captured by the spatially barcoded oligonucleotides on the Visium slide, an image is taken using the CytAssist instrument and superimposed onto the capture area which permits later H&E image alignment relative to the spatially resolved spots **(c)**. After the CytAssist transfer, the Visium slide is subjected to the standard workflow as per protocol CG000495 (10x Genomics). To generate the web summary output from a multiplexed capture area, manual selection of Visium spots covered by each tissue sample on Loupe Browser (V7 or higher; 10X Genomics) is required. This will allow the alignment of spatially resolved spots with H&E/CytAssist image. Our multiplexed array demonstrated higher gene counts in the epidermis/epidermal-dermal junction across all skin samples **(d)**. Clustering analysis confirms that the same cluster is assigned to the epidermis for all 17 samples **(e)**. For downstream analyses, each tissue within the capture area, as visualised on the Loupe Browser, is saved as individual file to enable independent analysis.

Here, we provide an example workflow with some key recommendations to enable tissue multiplexing. Prior to histopathological review, a fresh 4 µm section (5 µm thickness for our skin sections resulted in high level of tissue detachment during probe hybridisation and eosin wash) is acquired from the FFPE block with standard sectioning. At this stage, it is important to record the optimal water bath temperature by observing if the tissues disintegrate once floated. Tissue disintegration may be due to tissue block age, variable fixation times, tissue of origin, or the presence of abundant adipose cells. In our cohort, tissue ages ranged from 1-20+ years with water bath temperatures ranging from 30-40°C. To maximise the number of tissues that can fit into one Visium array, a dermatopathologist selects a single lesion of interest and the immediate perilesional area from each H&E-stained section. On the day of sectioning, blocks are submerged in UltraPure water (ThermoFisher Scientific, 10977015) and cooled for at least an hour on a cooling plate set at minus 4°C, to ensure the sample is rehydrated. An appropriately sized square is marked on the back of a positively charged slide (refer to 10x Genomics user guide #CG000520 for manufacturer recommendations) to guide section placement. During sectioning, the annotated H&E images guide selection of the correct area of interest. The microtome blade is repositioned and cleaned with RNase inhibitor solution (various providers) and 80% w/v ethanol for every tissue to avoid cross-contamination. To avoid potential RNA degradation with exposure to the air, the blocks are faced by discarding at least 12 µm from the surface of the block before a final 4 µm section is cut. Regions of interest are dissected from each section using a fresh scalpel blade (Medical and Surgical

Requisites, EU-210-1) by performing a dissection directly on the microtome stage, or forceps with an angled end (various providers) to perform dissection in the water bath. Excess paraffin is trimmed as much as possible, without damaging tissue integrity. The selected tissue piece is placed in the water bath at the previously determined suitable temperature for at least 30 seconds, before transferring it to the marked-up slide. Depending on the sample, the tissue might require a longer time in the water bath or, if it is old or under-fixed, the water bath incubation may be cut down to a few seconds. This process is repeated until all sections are placed as close as possible within the pre-drawn window(s) (**Figure 1a**). These curated slides are now ready to enter the standard Visium CytAssist workflow (10x Genomics CG000520, CG000495) (**Figure 1b-e**).

This tissue multiplexing workflow is a cost-effective means to increase sample size, whilst maintaining sample integrity and enabling statistically robust outcomes. These methods can be easily applied to most archival blocks collected from other tissue types.

## Data Availability

All data produced in the present study are available upon reasonable request to the authors

## Acknowledgements

The authors would like to thank 10x Genomics field application scientists Dr Catherine King, Dr James Fraser and Dr Gerry Ma for their technical support. The authors acknowledge the Translational Research Institute for providing an excellent research environment and core facilities that enabled this research. The authors particularly thank the histology and microscopy core members and Ms Emily Goy for their support.

## Notes

**Funding statement** This work was supported in part by the Office of the Assistant Secretary of Defense for Health Affairs through the Congressionally Directed Medical Research Programs Award No. (Melanoma Research Program-Discovery Award: W81XWH-21-MRP- DA) and the Merchant Charitable Foundation. Y-CK is supported by a UQ Graduate School Scholarships (UQGSS) and Research Training Program (RTP) funded by the Australian Government, KJL is supported by a National Health and Medical Research Council (NHMRC) Postgraduate Research Scholarship (GIN2013961) and a RTP scholarship. HPS was supported by a NHMRC Partnership Grant (1153046), MSS is supported by an NHMRC Investigator Award (2033440). KK is supported by a Cancer Council Grant (ACCR-0000095). The opinions, interpretations, conclusions, and recommendations are those of the author and are not necessarily endorsed by the Department of Defense. Funders had no role in the production of this manuscript.

**Conflict of interest:** disclosure HPS is a shareholder of MoleMap NZ Limited and e-derm consult GmbH, and undertakes regular teledermatological reporting for both companies. HPS provides medical consultant services for Canfield Scientific Inc. and First Derm by iDoc24 Inc The other authors have no conflicts of interest to disclose.

### Competing Interest Statement

: HPS is a shareholder of MoleMap NZ Limited and e-derm consult GmbH, and undertakes regular teledermatological reporting for both companies. HPS provides medical consultant services for Canfield Scientific Inc. and First Derm by iDoc24 Inc The other authors have no conflicts of interest to disclose.

### Funding Statement

This work was supported in part by the Office of the Assistant Secretary of Defense for Health Affairs through the Congressionally Directed Medical Research Programs Award No. (Melanoma Research Program-Discovery Award: W81XWH-21-MRP-DA) and the Merchant Charitable Foundation. Y-CK is supported by a UQ Graduate School Scholarships (UQGSS) and Research Training Program (RTP) funded by the Australian Government, KJL is supported by a National Health and Medical Research Council (NHMRC) Postgraduate Research Scholarship (GIN2013961) and a RTP scholarship. HPS was supported by a NHMRC Partnership Grant (1153046), MSS is supported by an NHMRC Investigator Award (2033440). KK is supported by a Cancer Council Grant (ACCR-0000095). The opinions, interpretations, conclusions, and recommendations are those of the author and are not necessarily endorsed by the Department of Defense. Funders had no role in the production of this manuscript.

### Author Declarations

The Metro South Health (Brisbane, Australia) Human Research Ethics Committee approved this study (approval number HREC/17/QPAH/816). The University of Queensland (Brisbane, Australia) Human Research Ethics Committees A and B confirmed the Metro South Heath ethics approval (University Of Queensland HREC approval number 2018000074). Participants gave written informed consent at the Princess Alexandra Hospital, Brisbane, Australia, according to the Declaration of Helsinki.

### Summary of Updates

This version of the manuscript has been revised to fix the typo in the abstract.

